# Multiplex PCR detection of enteric pathogens in a community-based birth cohort in Ecuador: comparison of xTAG-GPP and TaqMan array card assays

**DOI:** 10.1101/2024.10.10.24315212

**Authors:** Stuart Torres Ayala, Lesly Simbaña Vivanco, Nikolina Walas, Kelsey Jesser, Nicolette A. Zhou, Christine S. Fagnant-Sperati, Hadley R. Burroughs, Gwenyth O. Lee, Joseph N.S. Eisenberg, Gabriel Trueba, Karen Levy, Benjamin F. Arnold

## Abstract

We compared the performance of two multiplex platforms, Luminex xTAG Gastrointestinal Pathogen Panel^®^ and TaqMan Array Card, against a panel of 14 enteric pathogen targets in a community-based birth cohort in Ecuador. We found high levels of agreement and similar prevalence estimates across most pathogens.

## Introduction

Enteric pathogens account for a substantial burden of disease among children in low- and middle-income countries (1). Multiplex qPCR assays such as the Luminex xTAG Gastrointestinal Pathogen Panel® (GPP) and the TaqMan Array Card (TAC) enable efficient detection of pathogens in stool compared with single-pathogen PCR or qPCR testing and represent a major advance in enteric pathogen diagnostics (2,3). Multiplex assays were originally developed and have been used extensively in clinical settings for early infection diagnosis (4). Epidemiological studies are increasingly using these assays to characterize the burden of enteric infections, diarrheal etiology, and intervention impacts (5,6), and as the number of these population-based studies continues to grow, there is an increasing need to better understand the comparability of different multiplex assays. GPP is a commercial assay that screens for 15 pathogens while the TAC assay is customized by individual labs and, for this project, included 30 pathogens. We compared the performance of the two multiplex assays, GPP and TAC, against a panel of 14 overlapping viral, bacterial, and protozoan enteric pathogen targets in a community-based birth cohort in Ecuador, a high transmission setting.

## Methods

### Study Design

ECoMiD is an ongoing longitudinal birth cohort study based in Esmeraldas Province in northern-coastal Ecuador (7). The protocol was reviewed and approved by institutional review boards at University of Washington (#STUDY00014270), Emory University (#IRB00101202), Universidad San Francisco de Quito (#2018–022M), and University of California, San Francisco (#21-33932), and all participants provided informed consent, with re-consent for each stool sample collection. The study has enrolled 521 children from communities across a rural-urban gradient and collected periodic stool samples from study subjects from ages one week to 24 months. To potentially increase the efficiency of the assay comparison, we considered samples that had been analyzed by the TAC assay, run earlier in the study, and had a positive result for at least one pathogen in the GPP assay (n=485 samples). We then selected a random sample stratified by age (6, 12, 18 months) and location (rural accessible by river, rural accessible by road, intermediate, and urban) to be representative of the cohort (n=156, 13 per stratum). We estimated that 156 samples would provide 80% power to determine a difference between assays per target using McNemar’s test assuming a 5% alpha and sensitivities of 95.8% (TAC) and 89.6% (GPP), with conditional sensitivity of 98.9% for TAC given a positive by GPP (8) based on TAC and GPP parameters for rotavirus in clinical samples (2).

### Laboratory Methods

Stool samples were collected by caregivers in a small, insulated container and field staff collected samples within 1 hour of sample production if the sample was not refrigerated, or within 3 hours if the sample was refrigerated and stored in –196 ^°^C portable liquid nitrogen tanks. Samples were transported monthly to the Universidad San Francisco de Quito for long-term storage at –80 ^°^C.

Nucleic acids from stool samples (180-220 mg) were extracted using Qiagen QIAamp Fast DNA Stool Mini Kit (QIAGEN, Germantown, MD) into a proprietary elution buffer, with an added bead beating step during sample lysis (Jesser et al. *in review*). During the extraction process MS2 and PhHV were added as an external control assessment of extraction and amplification efficiency. ZymoBIOMICS Spike-in Controls (Zymo Research) were used as positive controls. Extracted DNA was aliquoted and stored at –80 ^°^C.

For GPP testing samples were amplified and hybridized according to the Luminex xTAG GPP kit protocol. xTAG^®^ RNAse-free water was used as a negative control and three stocks of known pathogen DNA (ZeptoMetrix NATtrol™ GI Verification Panel 2) were used as positive controls. GPP gene targets are proprietary but median fluorescence intensity (MFI) values used to determine positivity are available (**Supplemental Table 1**).

For TAC testing, extracted nucleic acids from stool samples (20 µL) were combined with AgPath-ID One-Step RT-PCR master mix (50 µL) (Applied Biosystems, Waltham, MA), AgPath-ID One-Step RT-PCR enzyme (4 µL) (Applied Biosystems), and nuclease-free water (26 µL) (Applied Biosystems) and analyzed for pathogen gene targets using TAC (ThermoFisher Scientific, Waltham, MA) (**Supplemental Table 2**) with the following cycling conditions: 45°C for 20 minutes, 95°C for 10 minutes, then 40 cycles of 95°C for 15 seconds and 60°C for 1 minute on a QuantStudio 7 Flex instrument (ThermoFisher Scientific). Positive controls included PhHV, MS2 and the pan *E. coli* gene target *uidA* as well as customized plasmids expressing all known assay targets (ThermoFisher Scientific and Azenta Life Sciences, South Plainfield, NJ). Nuclease-free water was used as a no template control on each card. Samples with cycle threshold (Ct) value ≤ 35 for any of the gene targets for a pathogen were classified as positive.

### Statistical Methods

We estimated pathogen target prevalence and agreement with exact, binomial 95% confidence intervals for the 14 targets. Agreement of pathogen target-level results between the two assays was assessed using McNemar’s test and Cohen’s kappa (9). Tests did not adjust for multiple comparisons. We examined MFI and Ct values for samples with discordant TAC and GPP results, positive by one assay and negative by the other, to determine if discordance was more likely with lower quantity of sample DNA detected. Analyses were conducted using R (v4.4.0; R Core Team 2024).

## Results

Two selected samples failed on the GPP assay, so the analysis included 154 samples. Overall, infection prevalence was similar between assays (**Figure 1**) and agreement was >85% for 13 of 14 pathogen targets (**Table 1**). There were differences in detection between TAC and GPP assays for five targets (McNemar’s P<0.05), with higher prevalence by TAC for rotavirus, *Campylobacter* spp., and ST-ETEC, and higher prevalence by GPP for *Shigella* spp., and *Salmonella* spp.. Accounting for agreement due to chance, six targets differed with a kappa coefficient below 0.6 (**Table 1**), however kappa statistics are influenced by outcome prevalence so comparison between pathogens should be made with caution given the wide range of prevalence observed (10). There was very poor agreement between assays for *Salmonella*, where the GPP assay classified 81% of samples as positive while the TAC assay classified 8% positive (**Figure 1, Table 1**). The rank order of prevalence was similar between assays with the exception of rotavirus, ST-ETEC, and *Salmonella*. Across targets, discordance between assays was more likely for pathogens with MFI values just over the positivity cutoff for GPP (for GPP+, TAC–, **Supplemental Figure 1**) or Ct value just below 35 for TAC (for TAC+, GPP–, **Supplemental Figure 2**).

**Table 1:**
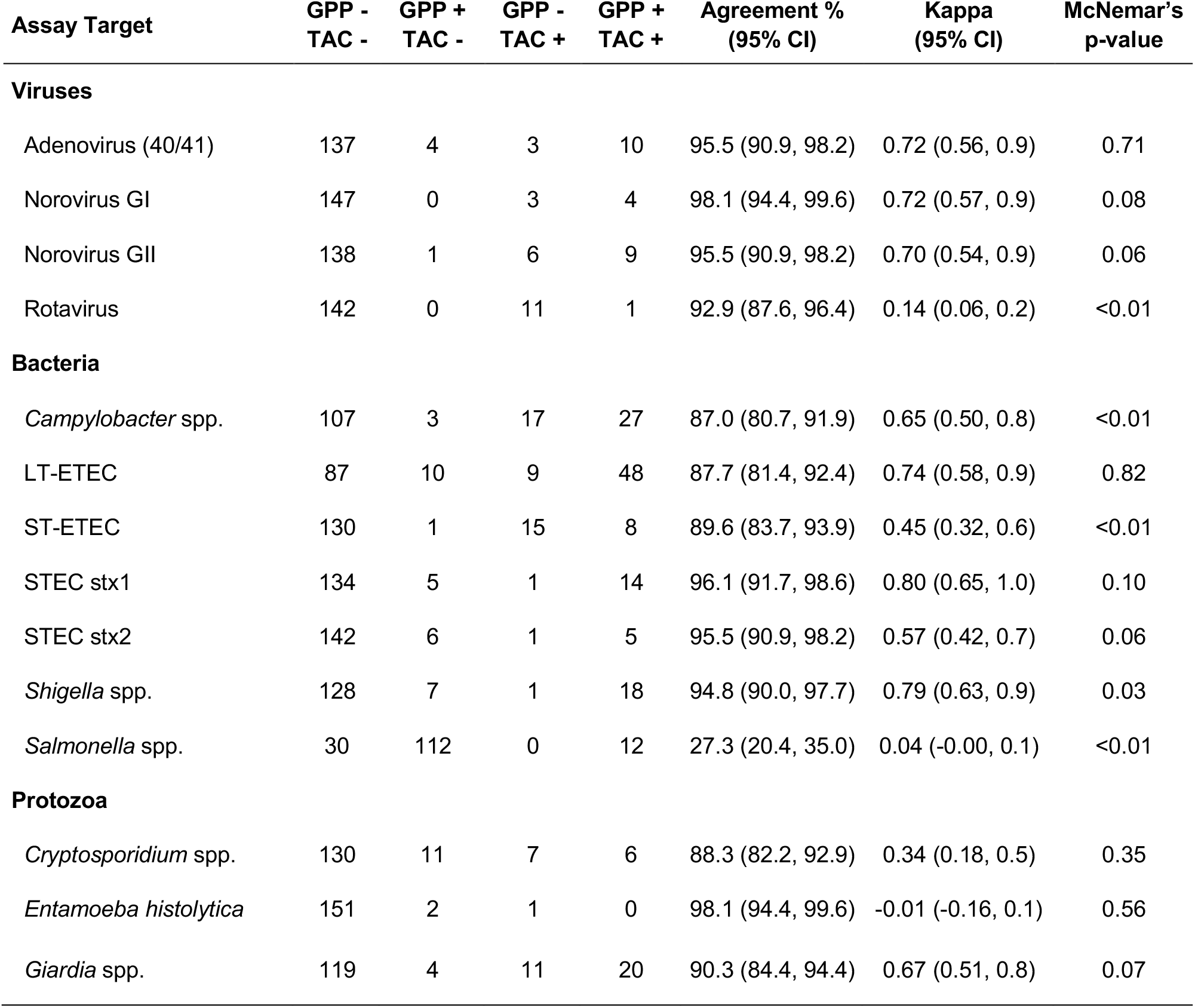
Summary of multiplex PCR test results for Luminex xTAG Gastrointestinal Pathogen Panel (GPP) and TaqMan Array Card (TAC). Test results from 154 samples measured among children at ages 6 to 18 months in Esmeraldas Province, Ecuador, 2022-2023. Test results are summarized by whether they were positive (+) or negative (–) by GPP and TAC. Methods include details on estimation of agreement, Cohen’s Kappa, and McNemar’s test for differences between assays. Created with script: https://osf.io/4dteq.

| Assay Target | GPP -<br>TAC - | GPP +<br>TAC - | GPP -<br>TAC + | GPP +<br>TAC + | Agreement %<br>(95% CI) | Kappa<br>(95% CI) | McNemar's<br>p-value |
| --- | --- | --- | --- | --- | --- | --- | --- |
| **Viruses** |  |  |  |  |  |  |  |
| Adenovirus (40/41) | 137 | 4 | 3 | 10 | 95.5 (90.9, 98.2) | 0.72 (0.56, 0.9) | 0.71 |
| Norovirus GI | 147 | 0 | 3 | 4 | 98.1 (94.4, 99.6) | 0.72 (0.57, 0.9) | 0.08 |
| Norovirus GII | 138 | 1 | 6 | 9 | 95.5 (90.9, 98.2) | 0.70 (0.54, 0.9) | 0.06 |
| Rotavirus | 142 | 0 | 11 | 1 | 92.9 (87.6, 96.4) | 0.14 (0.06, 0.2) | <0.01 |
| **Bacteria** |  |  |  |  |  |  |  |
| *Campylobacter* spp. | 107 | 3 | 17 | 27 | 87.0 (80.7, 91.9) | 0.65 (0.50, 0.8) | <0.01 |
| LT-ETEC | 87 | 10 | 9 | 48 | 87.7 (81.4, 92.4) | 0.74 (0.58, 0.9) | 0.82 |
| ST-ETEC | 130 | 1 | 15 | 8 | 89.6 (83.7, 93.9) | 0.45 (0.32, 0.6) | <0.01 |
| STEC stx1 | 134 | 5 | 1 | 14 | 96.1 (91.7, 98.6) | 0.80 (0.65, 1.0) | 0.10 |
| STEC stx2 | 142 | 6 | 1 | 5 | 95.5 (90.9, 98.2) | 0.57 (0.42, 0.7) | 0.06 |
| *Shigella* spp. | 128 | 7 | 1 | 18 | 94.8 (90.0, 97.7) | 0.79 (0.63, 0.9) | 0.03 |
| *Salmonella* spp. | 30 | 112 | 0 | 12 | 27.3 (20.4, 35.0) | 0.04 (-0.00, 0.1) | <0.01 |
| **Protozoa** |  |  |  |  |  |  |  |
| *Cryptosporidium* spp. | 130 | 11 | 7 | 6 | 88.3 (82.2, 92.9) | 0.34 (0.18, 0.5) | 0.35 |
| *Entamoeba histolytica* | 151 | 2 | 1 | 0 | 98.1 (94.4, 99.6) | -0.01 (-0.16, 0.1) | 0.56 |
| *Giardia* spp. | 119 | 4 | 11 | 20 | 90.3 (84.4, 94.4) | 0.67 (0.51, 0.8) | 0.07 |

**Figure 1:**
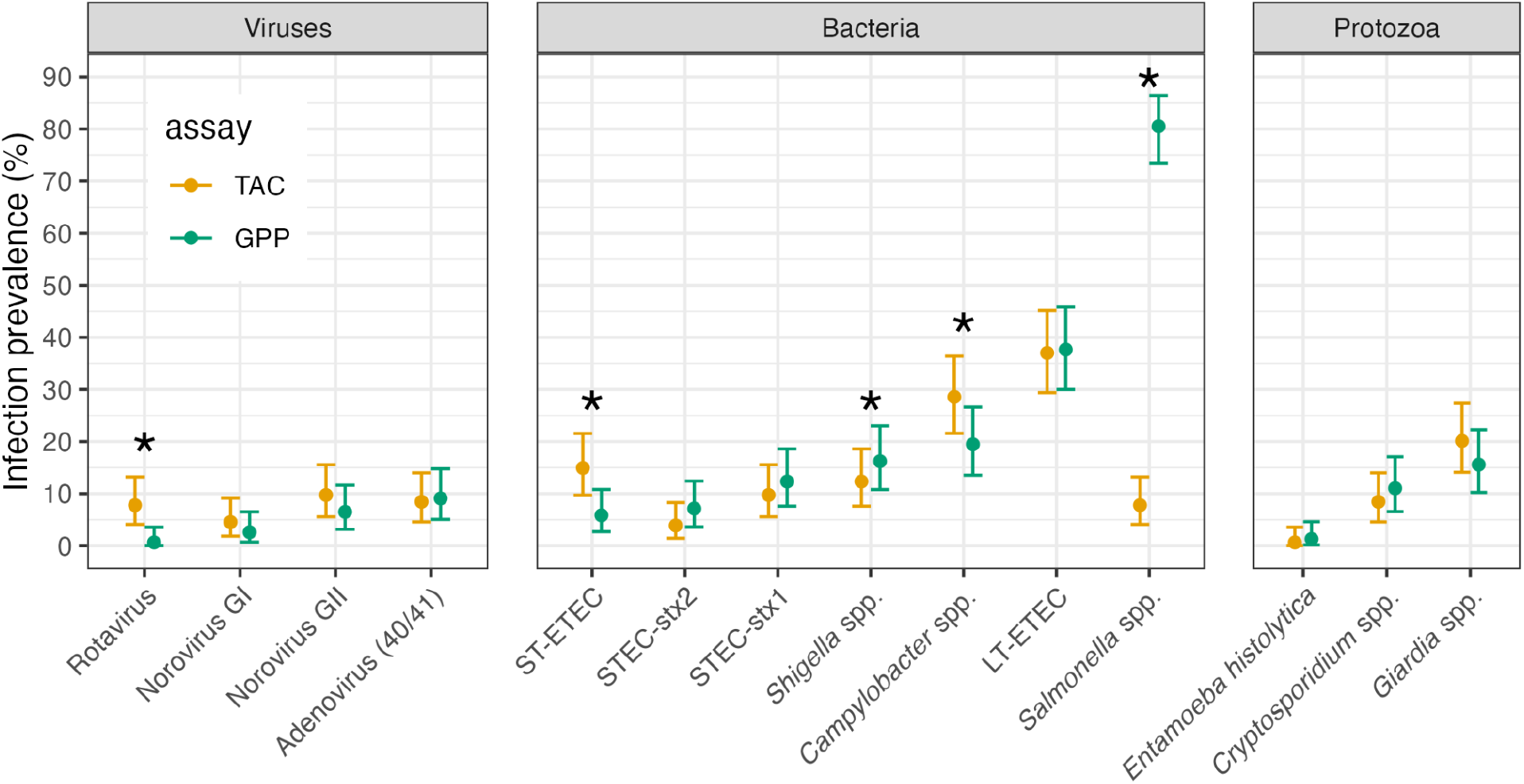
Infection prevalence for 14 enteric pathogens measured by Luminex xTAG Gastrointestinal Panel (GPP) and TaqMan Array Card (TAC) assays. Analysis includes 154 samples from children ages 6 to 18 months in Esmeraldas Province, Ecuador 2022-2023. An asterisk indicates McNemar’s P<0.05 for difference between the two assays. Supplemental Table 3 includes numerical estimates. Created with script: https://osf.io/4dteq.

## Discussion

Prior diagnostic comparison studies of the GPP and TAC assays have focused on tests of diarrheal samples in clinical settings, and found that the assays were broadly comparable and had good test performance as clinical diagnostics (2,6). This study aimed to evaluate the assay performance using community-based samples from young children and found the two assays were broadly comparable.

Consistent negative and positive controls on all GPP plates ruled out lab contamination as an explanation for the poor agreement between assays for *Salmonella*. Previous studies have noted high rates of *Salmonella* false positives by GPP (11,12) and at least one large-scale study excluded GPP *Salmonella* results on this basis (6). The discrepancy between GPP and TAC may result from differences in the oligonucleotide primers for the pathogen targets used for *Salmonella*.

This study had limitations. First, GPP uses proprietary target sequences — although we assume that differences between assay target sequences was an important underlying cause for larger discrepancies, such as for *Salmonella* and rotavirus, we could only infer this through examination of MFI and Ct values (**Supplemental Figures 1, 2**). Because we had no gold standard measure of infection across the 14 pathogens, we focused on agreement between the TAC and GPP assays but were unable to estimate their diagnostic characteristics, such as sensitivity and specificity. We focused on pathogen-specific comparisons between assays and did not assess co-infections or number of pathogens detected, which could be of interest in high transmission settings. We intentionally over-sampled stools that were positive by TAC to at least one target on the GPP assay to increase power for the comparison, but our sampling approach could inflate estimates of prevalence. Finally, we did not consider diarrhea symptoms in this analysis, but results should be representative of pediatric samples (both symptomatic and asymptomatic) in a high transmission setting.

Despite these caveats, this study had many strengths. We tested samples collected in a community-based cohort, with children enrolled across an urban-rural gradient at the ages when enteric pathogen burden is highest. The assays included pathogens thought to be major causes of diarrheal disease burden in lower resource settings (5), and we observed a broad range of pathogen prevalence in this study. The results thus should inform similar epidemiologic field studies.

## Conclusion

This comparative analysis provides important guidance on comparing data from TAC and GPP assays in non-clinical, pediatric samples for both within and across cohort analyses.

## Footnote information

### Funding

This work was funded by the National Institutes of Health (R01A137679 to KL and JNSE, R01AI162867 to BFA).

### Data availability

Data and replication files are available through the Open Science Framework: https://osf.io/jh64t/.

### Conference presentation

The results will be presented at the 2024 meeting of the American Society of Tropical Medicine and Hygiene, Oral Presentation Abstract #8373, November 17, 2024, New Orleans, LA, USA.

## Supplementary Information

**Supplemental Table 1:** Luminex Gastrointestinal Pathogen Panel (GPP) assay targets and corresponding median fluorescence intensity (MFI) thresholds for positivity.

| Analyte | Threshold for positivity (MFI) |
| --- | --- |
| Adenovirus 40/41 | ≥ 150 |
| *Camplobacter* | ≥ 150 |
| *C. difficile* Probe-1 | ≥ 150 |
| *C. difficile* Probe-2 | ≥ 150 |
| *Cryptosporidium* | ≥ 250 |
| *E. coli* O157 | ≥ 150 |
| *E. histolytica* | ≥ 250 |
| ETEC probe-1 | ≥ 200 |
| ETEC probe-2 | ≥ 200 |
| *Giardia* | ≥ 250 |
| Norovirus Probe-1 | ≥ 200 |
| Norovirus Probe-2 | ≥ 350 |
| Rotavirus A | ≥ 150 |
| *Salmonella* Probe-1 | ≥ 100,000 (POS), < 300 (NEG) |
| *Salmonella* Probe-2 | ≥ 200 |
| STEC Probe-1 | ≥ 150 |
| STEC Probe-2 | ≥ 150 |
| *Shigella* | ≥ 150 |
| *V. cholerae* | ≥ 150 |

**Supplemental Table 2:**
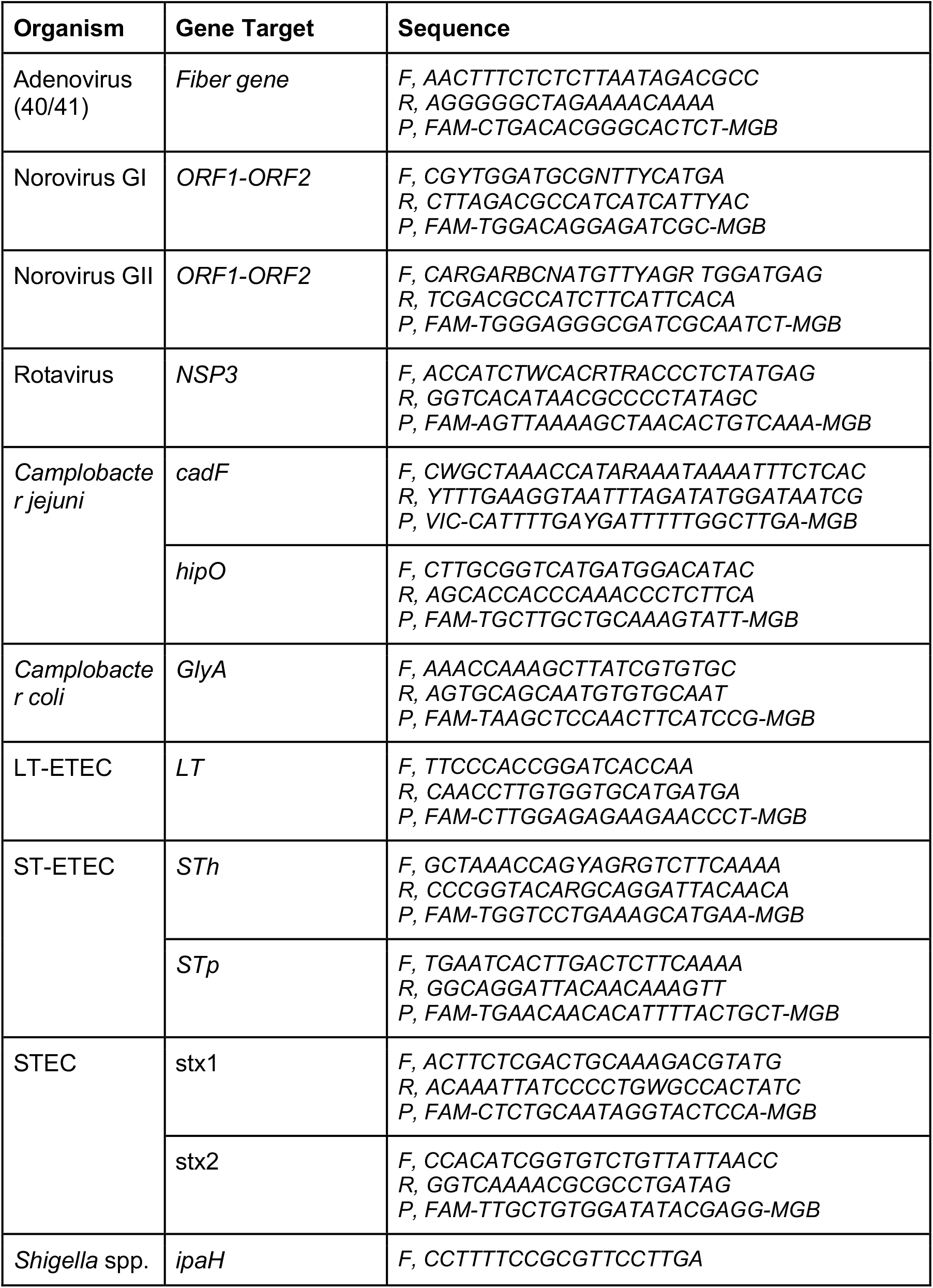
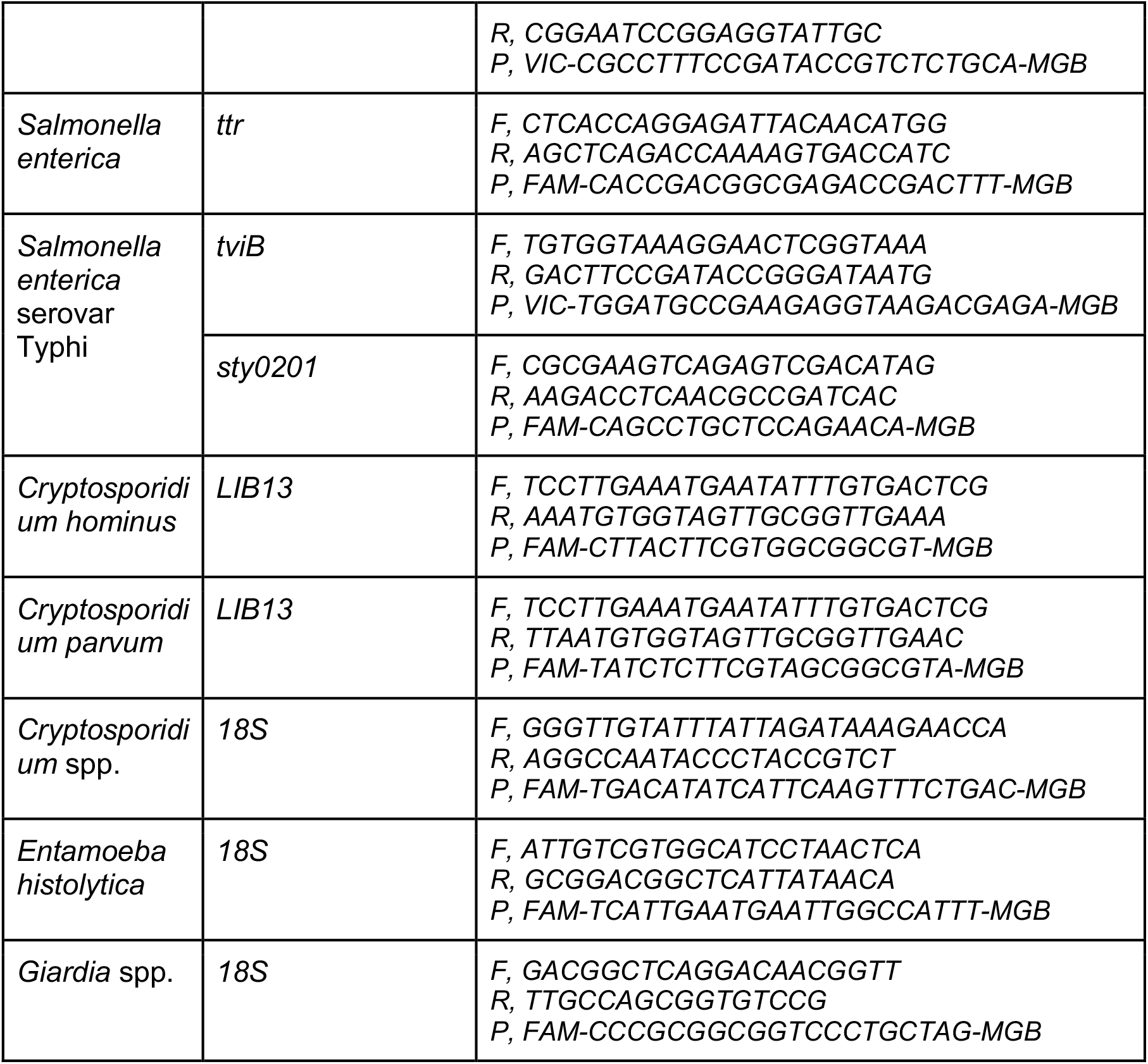
TaqMan Array Card (TAC) assay gene targets and corresponding forward (F) and reverse (R) primers and probe (P) sequences.

**Supplemental Table 3:** Infection prevalence for 14 enteric pathogens measured by Luminex xTAG Gastrointestinal Panel (GPP) and TaqMan Array Card (TAC) assays. Stool samples were tested from children at ages 6, 12, and 18 months old in Esmeraldas Province, Ecuador, 2022-2023. Created with script: https://osf.io/4dteq.

| GPP Target | N | GPP pos | GPP Prev (95% CI) | TAC pos | TAC Prev (95% CI) |
| --- | --- | --- | --- | --- | --- |
| **Viruses** |  |  |  |  |  |
| Adenovirus_40_41 | 154 | 14 | 9.1 (5.1, 14.8) | 13 | 8.4 (4.6, 14.0) |
| Norovirus_GI | 154 | 4 | 2.6 (0.7, 6.5) | 7 | 4.5 (1.8, 9.1) |
| Norovirus_GII | 154 | 10 | 6.5 (3.2, 11.6) | 15 | 9.7 (5.6, 15.6) |
| Rotavirus_A | 154 | 1 | 0.6 (0.0, 3.6) | 12 | 7.8 (4.1, 13.2) |
| **Bacteria** |  |  |  |  |  |
| Campylobacter | 154 | 30 | 19.5 (13.5, 26.6) | 44 | 28.6 (21.6, 36.4) |
| ETEC_LT | 154 | 58 | 37.7 (30.0, 45.8) | 57 | 37.0 (29.4, 45.2) |
| ETEC_ST | 154 | 9 | 5.8 (2.7, 10.8) | 23 | 14.9 (9.7, 21.6) |
| STEC_stx1 | 154 | 19 | 12.3 (7.6, 18.6) | 15 | 9.7 (5.6, 15.6) |
| STEC_stx2 | 154 | 11 | 7.1 (3.6, 12.4) | 6 | 3.9 (1.4, 8.3) |
| Shigella | 154 | 25 | 16.2 (10.8, 23.0) | 19 | 12.3 (7.6, 18.6) |
| Salmonella | 154 | 124 | 80.5 (73.4, 86.5) | 12 | 7.8 (4.1, 13.2) |
| **Protozoa** |  |  |  |  |  |
| Cryptosporidium | 154 | 17 | 11.0 (6.6, 17.1) | 13 | 8.4 (4.6, 14.0) |
| Entamoeba_histolytica | 154 | 2 | 1.3 (0.2, 4.6) | 1 | 0.6 (0.0, 3.6) |
| Giardia | 154 | 24 | 15.6 (10.2, 22.3) | 31 | 20.1 (14.1, 27.3) |

**Supplemental Figure 1:**
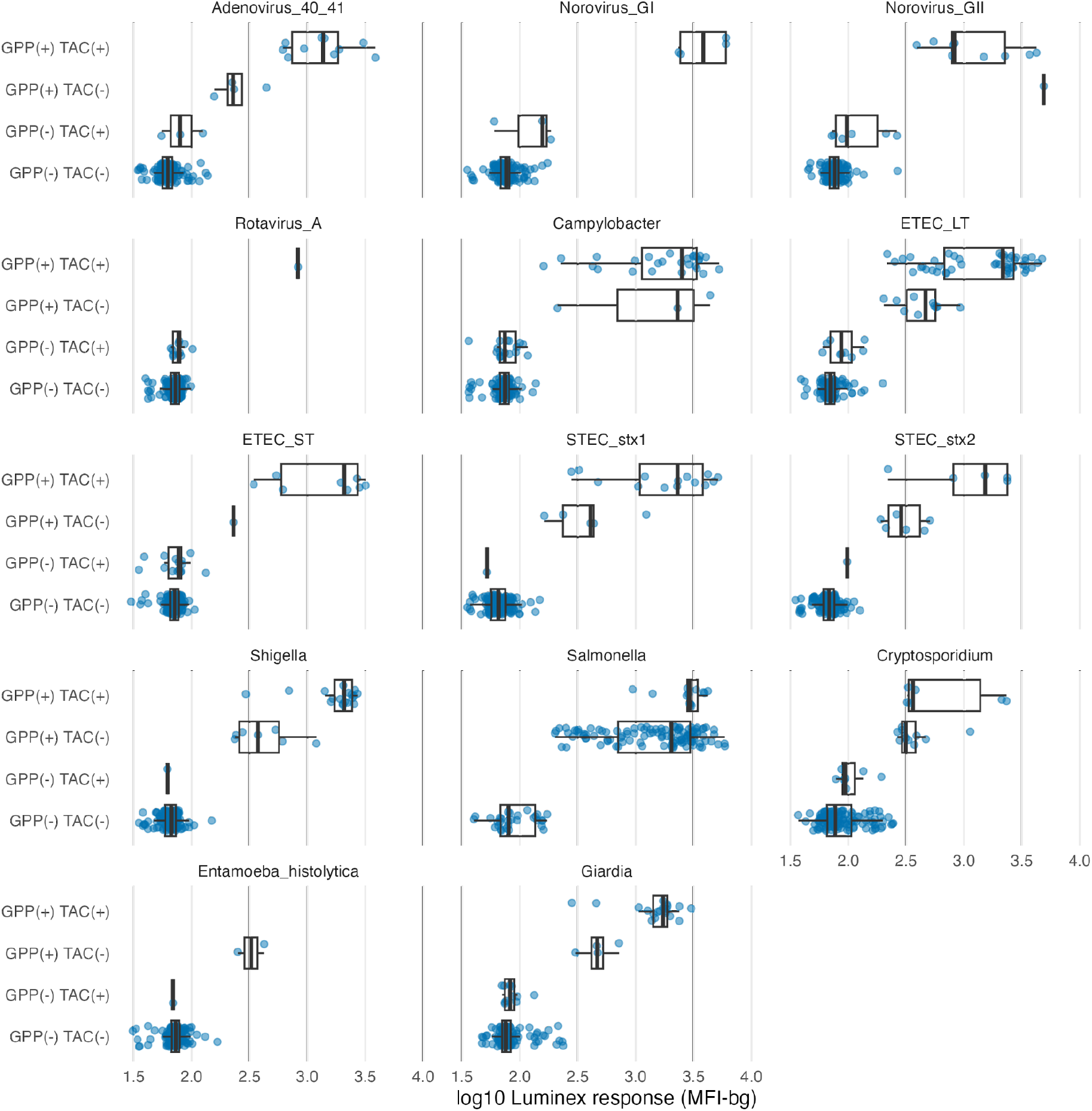
Median fluorescence intensity (MFI) values for pathogen targets detected by the Luminex GPP Assay. Results are categorized according to TaqMan Array Card (TAC) and Luminex xTAG Gastrointestinal Pathogen Panel (GPP) assay sample results, for positive (+) and negative (–) detection for each target. Created with script: https://osf.io/hfv5r.

**Supplemental Figure 2:**
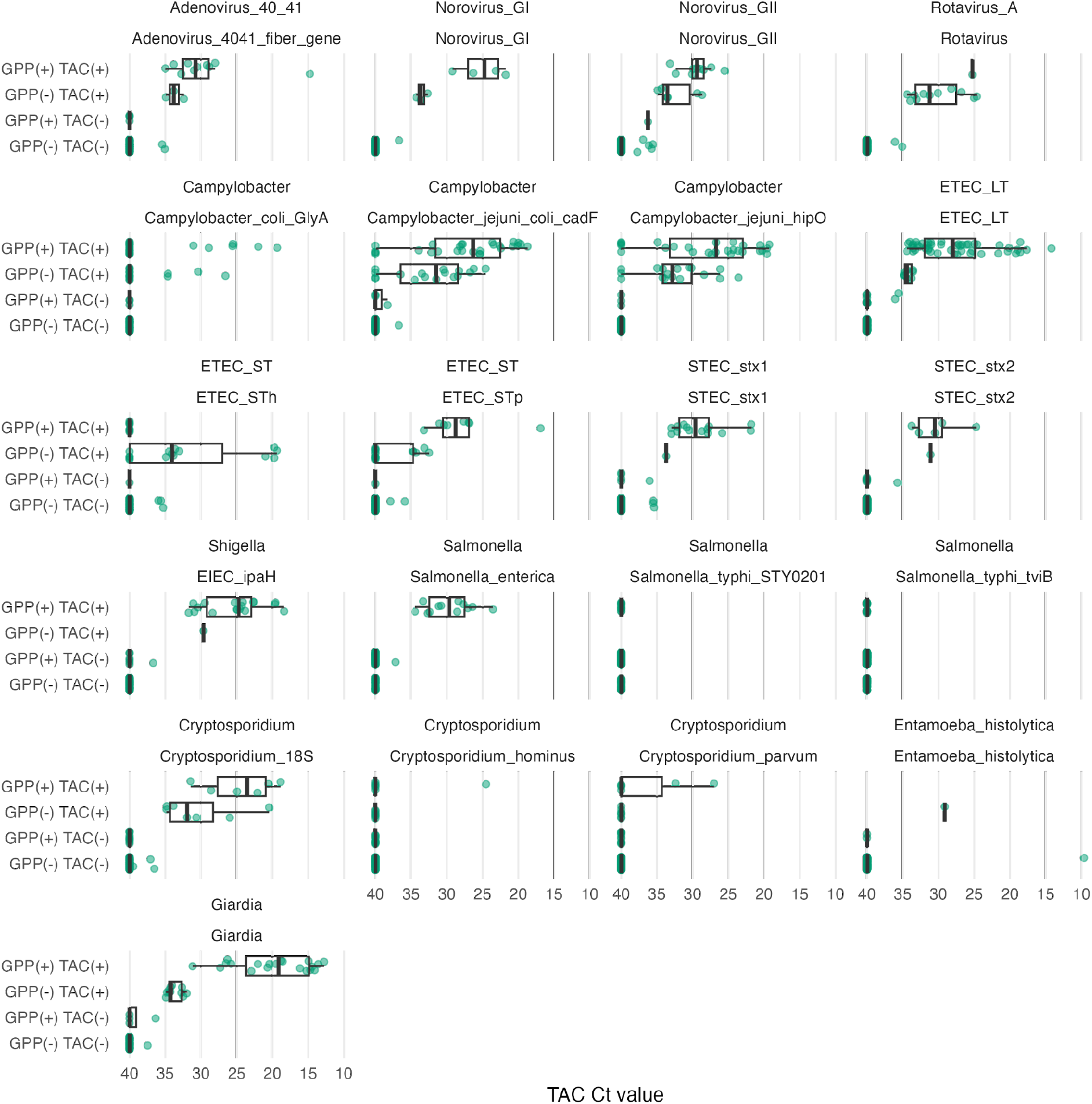
Cycle threshold (Ct) values for pathogen associated gene targets detected by the TAC Assay. Multiple gene targets were used for some enteric pathogens in the TaqMan Array Card (TAC) panel. In each comparison, the top row label identifies the Luminex xTAG Gastrointestinal Pathogen Panel (GPP) target, and the second row identifies the TAC target. Results are categorized according to TAC and GPP assay sample results, for positive (+) and negative (–) detection for each GPP target. Created with script: https://osf.io/hfv5r.

